# Multifocal breast cancers are more prevalent in *BRCA2* versus *BRCA1* mutation carriers

**DOI:** 10.1101/19006478

**Authors:** Alan D McCrorie, Susannah Ashfield, Aislinn Begley, Colin Mcilmunn, Patrick J. Morrison, Clinton Boyd, Bryony Eccles, Stephanie Greville-Heygate, Ellen R Copson, Ramsey I. Cutress, Diana M Eccles, Kienan I. Savage, Stuart A McIntosh

## Abstract

Multifocal/multicentric breast cancer is generally considered to be where two or more breast tumours are present within the same breast, and is seen in ∼10% of breast cancer cases. This study investigates the prevalence of multifocality/multicentricity in a cohort of *BRCA1/2* mutation carriers with breast cancer from Northern Ireland via cross-sectional analysis. Data from 211 women with *BRCA1/2* mutations (*BRCA1* - 91), (*BRCA2* - 120), with breast cancer were collected including age, tumour focality, size, type, grade, and receptor profile. The prevalence of multifocality/multicentricity within this group was 25%, but within subgroups, prevalence amongst *BRCA2* carriers was more than double that of *BRCA1* carriers (p=0.001). Women affected by multifocal/multicentric tumours had proportionately higher oestrogen receptor positivity (p=0.001) and lower triple negativity (p=0.004). These observations are likely to be driven by the higher BRCA2 mutation prevalence observed within this cohort. Odds of a *BRCA2* carrier developing multifocal/multicentric cancer were almost four-fold higher than a *BRCA1* carrier (OR: 3.71, CI: 1.77-7.78, p=0.001). These findings were subsequently validated in a second, large independent cohort of patients with *BRCA*-associated breast cancers from a UK-wide multicentre study. This confirmed a significantly higher prevalence of multifocal/multicentric tumours amongst *BRCA2* mutation carriers compared with *BRCA1* mutation carriers. This has important implications for clinicians involved in the treatment of BRCA2-associated breast cancer, both in the diagnostic process, in ensuring that tumour focality is adequately assessed to facilitate treatment decision-making, and for breast surgeons, particularly if breast conserving surgery is being considered as a treatment option for these patients.

## Introduction

A large meta-analysis of 22 studies, including over 67,000 women, estimated prevalence of multifocal breast cancer to be 9.5%[1]. Although multifocality does not appear to be an independent predictor of outcome in breast cancer, the sum of the invasive foci in multifocal disease may be associated with reduced disease-free survival, when compared with unifocal tumours[2–4]. Moreover, treatments offered for multifocal breast cancer vary widely, with some women undergoing multiple breast conserving procedures, and others mastectomy, with no clear treatment guidelines[5].

Historically, the definitions of multifocal (MF) and multicentric (MC) breast cancer have varied. Multifocal cancers have been defined as two or more distinct invasive breast carcinomas within the same breast quadrant, whereas multicentric disease has been defined as separate tumours in different breast quadrants. Studies have suggested that in cases of both MF and MC disease, tumours may either share or be of independent clonal origin [6–9]. Furthermore, published data suggests that MF and MC disease may have different patterns of behaviour clinically [10]. Due to these conflicting definitions and the lack of clarity on whether these entities represent the same disease process, for the pragmatic purposes of this retrospective study and to avoid confusion, we have considered MF and MC tumours together.

BRCA1 and BRCA2 are tumour suppressor genes located on chromosomes 17 and 13 respectively. They encode proteins involved in the cellular DNA damage response pathway, particularly DNA double strand break repair[11]. Germline mutations in these genes predispose female carriers to a significantly increased risk of breast and ovarian cancer, with up to 80% lifetime risk of breast cancer. Given this elevated breast cancer risk, we hypothesised that these women may be more likely than non-mutation carriers to develop multifocal disease. Surprisingly, despite biological plausibility for the existence of an association between BRCA1/2 mutations and MF/MC tumours, at time of writing, there were no studies investigating this. Therefore, this study aimed to investigate the prevalence of MF/MC breast cancer in BRCA1/2 mutation carriers, with exploration of the clinicopathological characteristics of all tumours occurring in these patients.

## Materials and methods

Data from 252 women with a known pathogenic germline *BRCA1/2* mutation diagnosed with breast cancer (1994-2017) were retrospectively extracted from a database containing all known female *BRCA1/2* mutation carriers in Northern Ireland **(Figure 1)**. Information about histological tumour type and focality (unifocal or multifocal) was extracted from pathology records for 211 women, with 41 patients excluded due to missing focality information (n=30), or because of a diagnosis of DCIS without invasion (n=11). Additional clinicopathological data was collected, including age at initial cancer diagnosis, tumour grade and size, hormone receptor status, HER2 status, nodal involvement, presence/absence of other primary cancer. Outcome data was collected from electronic hospital records, and cause of death ascertained. Data was entered into Microsoft Excel® for stratification and calculation of prevalence. 23 randomly selected cases (10%) underwent review of the original diagnostic slides by an independent pathologist for validation of multifocality/multicentricity reporting.

**Figure 1:**
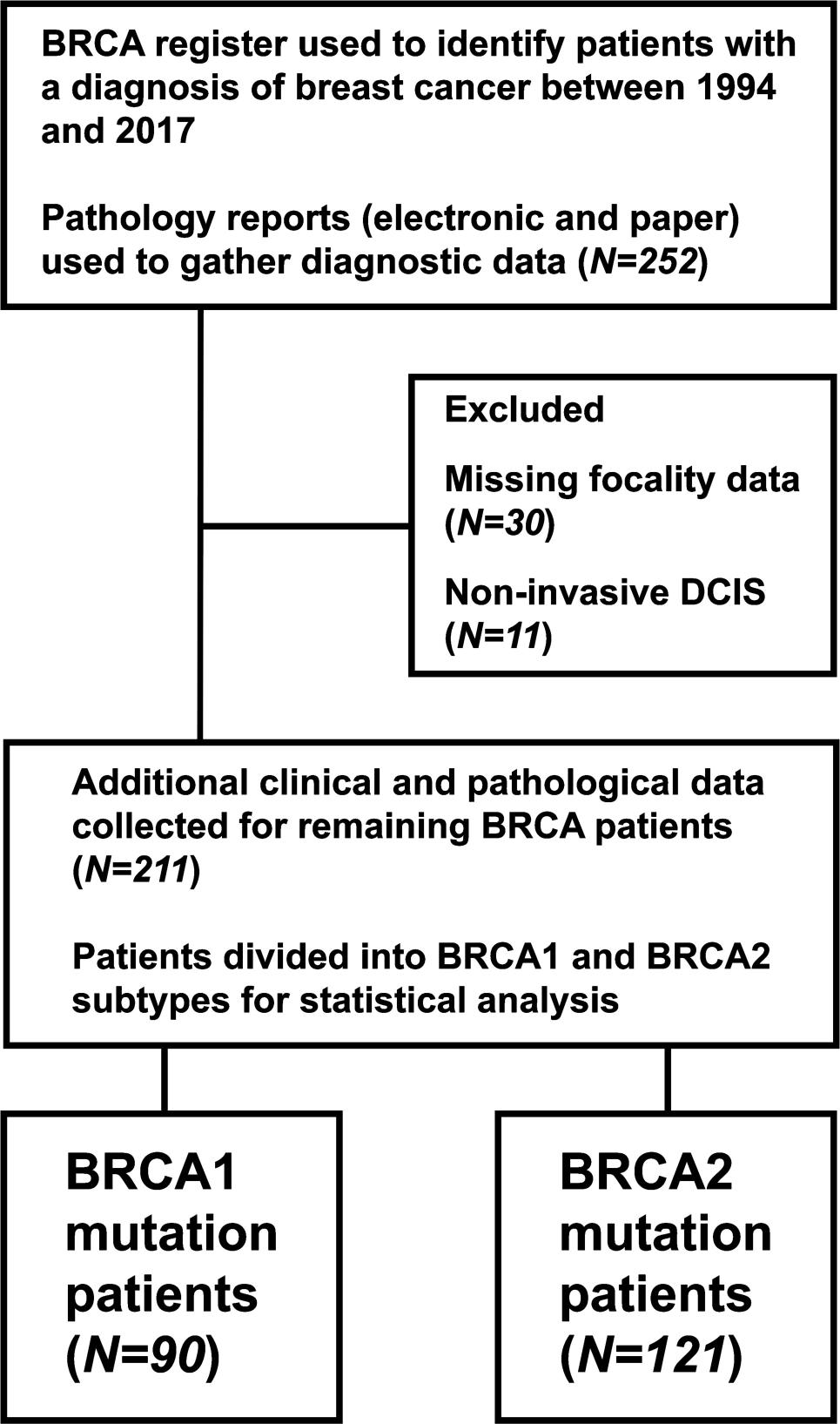
participant flow diagram showing inclusions/exclusions.

For validation of the findings in the Northern Ireland patient cohort, a second cohort of breast cancer patients with known germline BRCA1/2 mutations was identified. The POSH (Prospective Outcomes in Sporadic versus Hereditary breast cancer) study recruited young women (aged 18-40) diagnosed with primary breast cancer in the United Kingdom between 2000 and 2008 [12]. The study methodology (including genotyping methods) and outcomes have previously been reported [13]. Data on tumour focality/centricity in the POSH study patients was obtained from medical records from participating centres.

Data were analysed using SPSS®. Heterogeneity of clinicopathological characteristics between those diagnosed with unifocal disease and those diagnosed with MF/MC disease were compared using χ^2^. Mean age and tumour size between groups was compared using t-test. Binary logistic regression was performed to calculate the unadjusted odds ratio (OR) of developing multifocal disease in patients with *BRCA2*-associated breast cancer versus those with *BRCA1*-associated breast cancer. Thereafter, adjusted OR was calculated using a manually controlled backward stepwise elimination approach[14]. Potentially confounding variables with a biological association to breast cancer were entered into the regression model and sequentially removed until only those with statistical significance remained. Survival estimates were carried out using the Kaplan-Meier method. A p value of <0.05 indicates significance at the 95% confidence interval throughout. Institutional approval was granted by the Belfast Health and Social Care Trust (Ref: 5805). Ethical approval for the POSH study was granted in 2000 (MREC 00/6/69).

## Results

### Northern Ireland BRCA1/2 cohort

Of the 211 BRCA1/2 carriers diagnosed with breast cancer (with MF/MC information available) in Northern Ireland between 1994-2017 90 (42.7%) women had a *BRCA1* mutation and 121 (57.3%) a *BRCA2* mutation. Mean age at diagnosis was 45 years (range: 25-77 years) with a lower mean age at diagnosis for MF/MC tumours compared with unifocal tumours (43 vs. 46 years) (p=0.109). Mean tumour size was 24mm (range: 2-150mm) with no significant difference in mean size between the largest MF/MC tumour foci and unifocal tumours (24.8mm vs. 23.2mm) (p=0.587). There were 52 diagnoses of MF/MC disease and 159 diagnoses of unifocal disease. Prevalence of MF/MC disease was 13.3% in *BRCA1* mutation carriers and 33.1% in *BRCA2* mutation carriers. Therefore, prevalence of MF/MC disease in *BRCA2* carriers was 2.5-fold greater than *BRCA1* carriers (p=0.001). Clinicopathological findings are documented in **Table 1**. The majority of MF/MC and unifocal tumours were invasive ductal carcinomas (86.5% and 96.2% respectively), grade III (73.6% and 63.5% respectively), and HER2-negative (75.0% and 73.6% respectively). Additionally, *BRCA1/2* carriers with MF/MC disease were more likely to be oestrogen receptor positive than negative (75.0% vs. 45.9%) (p=0.001). Furthermore, it is known that invasive lobular carcinoma (ILC) is seen more frequently in BRCA2 than BRCA1 mutation carriers [15]. We therefore excluded the 9 cases of ILC in this cohort (7 BRCA2 mutation carriers and 2 BRCA1 mutation carriers), and repeated the analysis including invasive ductal carcinoma alone, showing that multifocality/multicentricity remained significantly higher in BRCA2 versus BRCA1 mutation carriers when cases of ILC were excluded (p=0.001).

**Table 1:** Clinical and pathological characteristics of *BRCA1/2* mutation carrier patients diagnosed with breast cancers in Northern Ireland between 1994-2017. *Pearson’s χ^2^ where p<0.05 indicates significance.

| Clinical and pathological features of breast cancers |  | Multifocality<br>N (%) | Unifocality<br>N (%) | p-value<br>(*) |
| --- | --- | --- | --- | --- |
| BRCA mutation | BRCA1 | 12 (13.3) | 78 (86.7) | 0.001 |
|  | BRCA2 | 40 (33.1) | 81 (66.9) |  |
| Age at first diagnosis | <40 years | 23 (32.9) | 47 (67.1) | 0.039 |
|  | ≥40 years | 29 (20.6) | 112 (79.4) |  |
| Tumour subtype | Invasive ductal | 45 (22.7) | 153 (77.3) | 0.011 |
|  | Invasive lobular | 6 (66.7) | 3 (33.3) |  |
|  | Other | 1 (25.0) | 3 (75.0) |  |
| Tumour grade | I | 3 (42.9) | 4 (57.1) | 0.460 |
|  | II | 15 (30.0) | 35 (70.0) |  |
|  | III | 33 (22.0) | 117 (78.0) |  |
|  | Unknown | 1 (25.0) | 3 (75.0) |  |
| Oestrogen receptor | Positive | 39 (34.8) | 73 (65.2) | 0.001 |
|  | Negative | 12 (12.9) | 81 (87.1) |  |
|  | Unknown | 1 (16.7) | 5 (83.3) |  |
| HER2 status | Positive | 3 (27.3) | 8 (72.7) | 0.933 |
|  | Negative | 39 (25.0) | 117 (75.0) |  |
|  | Unknown | 10 (22.7) | 34 (77.3) |  |
| Triple negativity | Yes | 7 (12.5) | 49 (87.5) | 0.004 |
|  | No | 30 (36.1) | 53 (63.9) |  |
|  | Unknown | 15 (20.8) | 57 (79.2) |  |
| Lymph node involvement | Yes | 26 (33.3) | 52 (66.7) | 0.072 |
|  | No | 25 (20.0) | 100 (80.0) |  |
|  | Unknown | 1 (12.5) | 7 (87.5) |  |
| Presence of other primary cancer | Yes | 9 (23.7) | 29 (76.3) | 0.957 |
|  | No | 42 (25.0) | 126 (75.0) |  |
|  | Unknown | 1 (20.0) | 4 (80.0) |  |

Of the 52 women diagnosed with MF/MC disease, 23.1% (n=12) had a *BRCA1* mutation and 76.9% (n=40) a *BRCA2* mutation. 50% (n=6) of women with a *BRCA1* mutation were oestrogen receptor positive whilst 82.5% (n=33) women with a *BRCA2* mutation were oestrogen receptor positive (p=0.039). See **Supplementary Table 1**. Unadjusted odds of breast cancer being MF/MC in *BRCA2* mutation carriers were 3.2 times greater than in *BRCA1* mutation carriers (CI:1.57–6.57, p=0.001). Age was found to be a significant confounding factor in logistic regression (CI:0.22–0.85, p=0.015). Therefore, after adjusting for age, odds of a *BRCA2* mutation carrier developing MF/MC breast cancer were 3.7-fold greater than in *BRCA1* mutation carriers (CI:1.77–7.78, p=0.001) (Table 3).

**Table 2:**
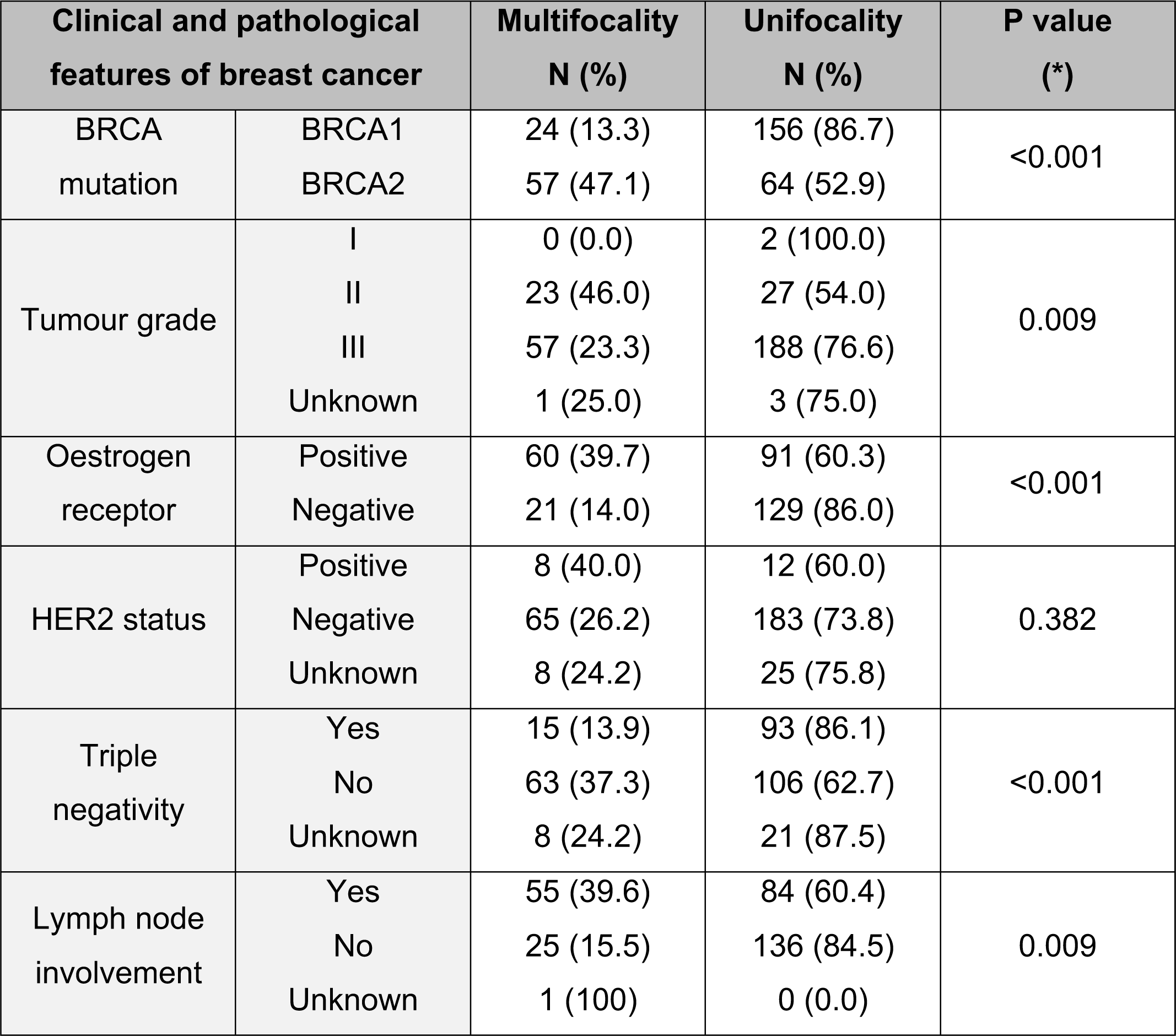
Clinical and pathological characteristics of BRCA1/2 mutant carrier patients diagnosed with breast cancers within POSH dataset (2000-2008). *Pearson’s χ^2^ where p<0.05 indicates significance.

**Table 3:**
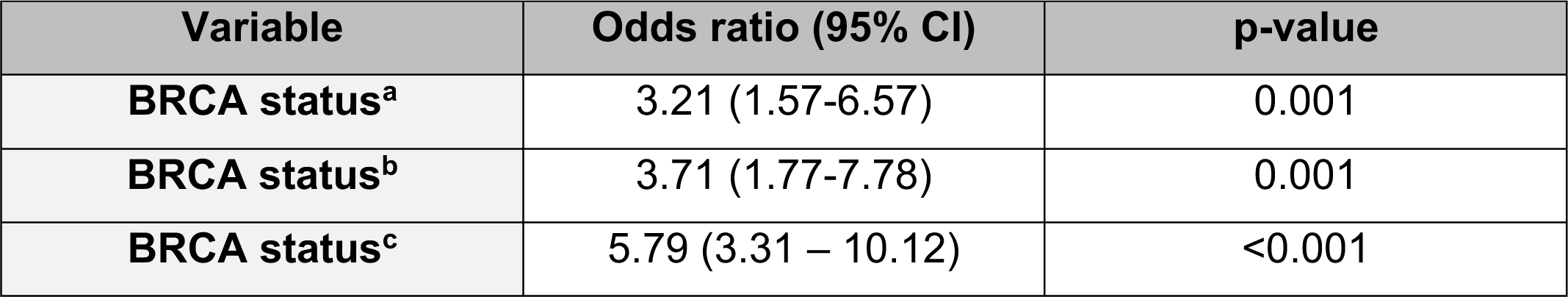
Odds of cancer being multifocal in patients with BRCA2 versus BRCA1 mutation, where (a) = unadjusted odds ratio in Northern Ireland cohort, (b) = adds ratio in Northern Ireland cohort adjusted for age (≥40 years versus < 40 years) and (c) = unadjusted adds ratio in POSH study cohort

| Variable | Odds ratio (95% CI) | p-value |
| --- | --- | --- |
| **BRCA status<sup>a</sup>** | 3.21 (1.57-6.57) | 0.001 |
| **BRCA status<sup>b</sup>** | 3.71 (1.77-7.78) | 0.001 |
| **BRCA status<sup>c</sup>** | 5.79 (3.31 – 10.12) | <0.001 |

At a median follow-up of 9.5 years for the cohort of Northern Irish patients, there was no breast cancer specific survival difference between women with MF/MC versus unifocal disease (log-rank p=0.617), and when adjusted for BRCA mutation status this remained non-significant (log-rank p=0.775)(Figure 2A). Similarly, there was no difference in survival between MF/MC or unifocal tumours in BRCA1 mutation carriers (Figure 2B), BRCA2 mutation carriers (Figure 2C), nor was there a difference in all-cause survival between women with unifocal versus MF/MC disease (Figure 2D).

**Figure 2:**
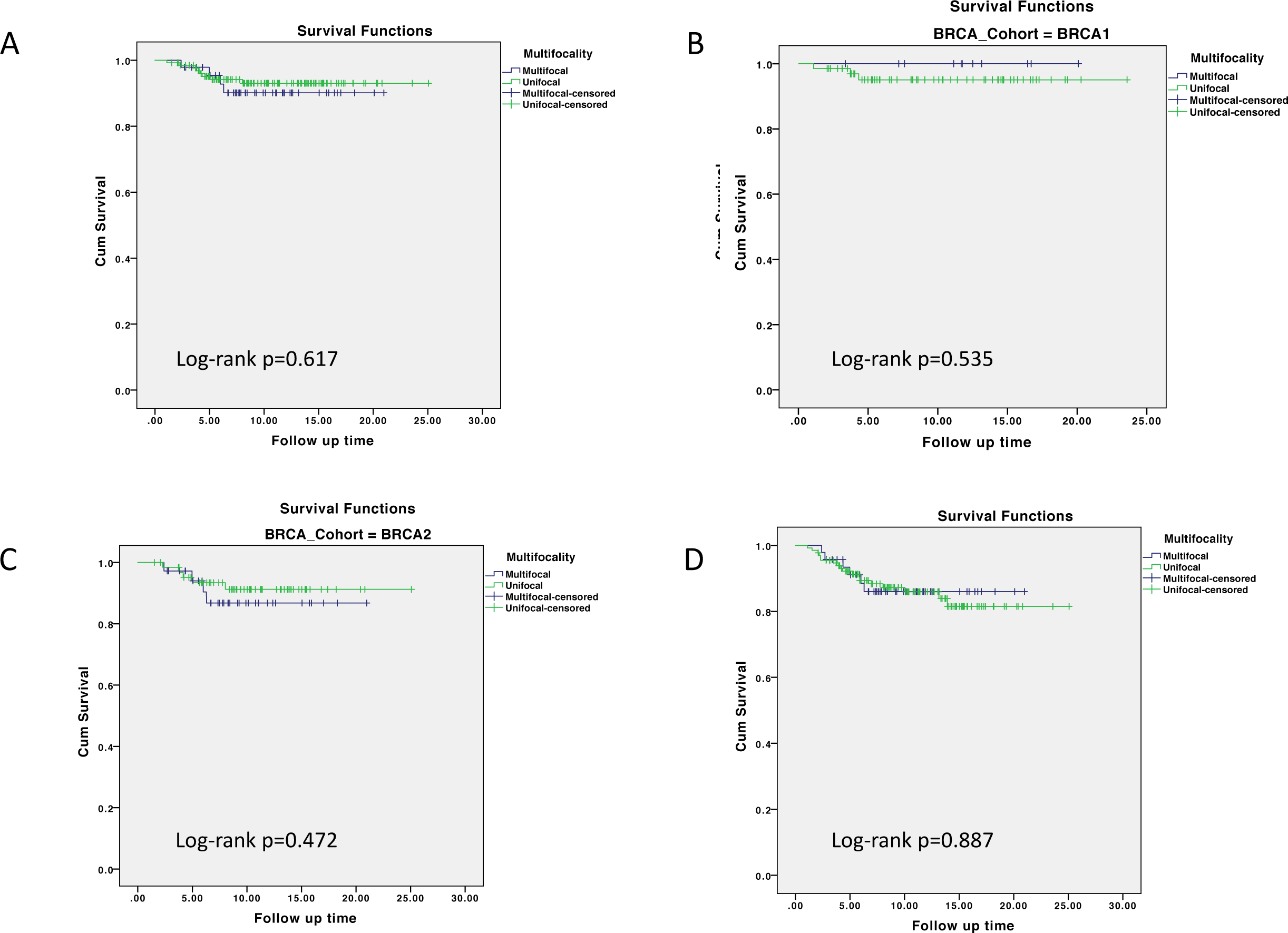
Survival outcomes in the Northern Ireland patient cohort. A=MF/MC disease *versus* unifocal disease, breast cancer specific mortality, all patients (*BRCA1/2*). B=MF/MC disease *versus* unifocal disease, breast cancer specific mortality in *BRCA1* mutation carriers. C=MF/MC disease *versus* unifocal disease, breast cancer specific survival in *BRCA2* mutation carriers. D=MF/MC *versus* unifocal disease, all -cause mortality in all patients (*BRCA1/2*).

### POSH study BRCA1/2 cohort

There were 338 germline BRCA mutation carriers in the POSH study breast cancer cohort; focality data was missing in 37 cases, leaving 180 women with a BRCA1 mutation and 121 with a BRCA2 mutation for analysis. There were 81 diagnoses of MF/MC disease and 220 diagnoses of unifocal disease. Clinicopathological findings in the POSH cohort are detailed in Table 2. Mean age of diagnosis was 34 years, with no difference seen in the age at diagnosis for MF/MC tumours versus unifocal tumours (35 versus 34 years). MF/MC breast cancer was identified in 26.9% of BRCA1/2 mutation carriers who developed breast cancer. Prevalence of MF/MC disease was 13.3% amongst BRCA1 mutation carriers diagnosed with breast cancer, and 47.1% amongst BRCA2 mutation carriers diagnosed with breast cancer. Therefore, prevalence of MF/MC disease in BRCA2 mutation carriers was 3.5-fold greater than in BRCA1 mutation carriers in this independent cohort of BRCA1/2 carriers (P<0.001).

BRCA1/2 mutation carriers with MF/MC disease were more likely to be oestrogen receptor positive (74.1%) than those with unifocal disease (41.4%). This difference in proportions was significant (P<0.001). Similarly, BRCA1/2 mutation carriers with MF/MC disease were less likely to be triple receptor negative (18.5%) compared to those with unifocal disease (42.3%). This was also significant (P<0.001). When data from women who developed MF/MC breast cancer was analysed in isolation, prevalence of oestrogen receptor positivity was 85% amongst BRCA2 mutation carriers but only 15% in BRCA1 mutation carriers. This difference in proportions was significant (p<0.001).

The unadjusted odds (in binary logistic regression analysis) of a breast cancer being MF/MC in BRCA2 mutation carriers was 5.8 times greater than in a BRCA1 mutation carrier who developed breast cancer (CI: 3.31-10.12) (p<0.001) (Table 3). Adjustment for oestrogen receptor status gave odds of a BRCA2 mutation carrier developing MF/MC breast cancer 4.2 times greater than a BRCA1 mutation carrier (CI: 2.12-8.19) (p<0.001). A similar reduction in the magnitude of the odds ratio was observed in our dataset when analysis was adjusted for oestrogen receptor status.

## Discussion

A systematic review of sporadic MF/MC breast cancer conducted by Vera-Badillo *et al* included twenty-two studies encompassing 67,557 women. This study calculated a prevalence of 9.5% amongst women with sporadic breast cancer (*BRCA* status unknown)[1]. It should also be noted that this meta-analysis only includes women with early stage breast cancer, and only includes studies which provided survival outcome data, so it is possible that this is not truly representative of the incidence of MF/MC disease in the general population. Nevertheless, and in contrast, the prevalence of multifocality/multicentricity in the Northern Ireland cohort of 211 *BRCA1/2* mutation carriers was 24.6%, more than double that reported by Vera-Badillo *et al* for sporadic breast cancer [1]. This finding is supported by a strikingly similar prevalence of 26.9% in the larger cohort of BRCA1/2 mutation carriers from the multicentre POSH study.

Our study found that prevalence of multifocality/multicentricity in *BRCA2* mutation carriers was at least double that in *BRCA1* mutation carriers in both reported patient cohorts – a finding mirrored in a small-scale study by Bergthorsson *et al[16]*. The odds of a woman who has developed breast cancer, exhibiting MF/MC disease, are over three times greater if she has a *BRCA2* mutation compared to a *BRCA1* mutation. This rises to an almost four-fold increase in odds of a *BRCA2* carrier developing MF/MC disease once the effect of age at diagnosis is taken into account.

Women diagnosed with MF/MC breast cancer were proportionately more likely to be oestrogen receptor positive and had a lower prevalence of triple receptor negativity in both the Northern Ireland and POSH study patients. These findings are in keeping with numerous studies documenting significantly higher rates of oestrogen receptor positivity amongst *BRCA2* carriers compared with *BRCA1* mutation carriers. Therefore, it is unlikely that oestrogen signalling itself drives MF/MC disease[17,18]. Indeed, the large-scale meta-analysis described earlier found no association between ER status and sporadic MF/MC breast cancer, suggesting that ER activity does not play a role in the specific development of MF/MC disease[1]. Similarly, although ILC is seen more commonly in *BRCA2* mutation carriers than in *BRCA1* carriers in the Northern Ireland patient cohort, the significant increase in prevalence of MF/MC disease in *BRCA2* carriers persists even when ILC cases are excluded from the analysis, suggesting that it is not the lobular phenotype which drives the increased prevalence of multifocality/multicentricity.

Precisely why *BRCA2* carriers are more likely to develop MF/MC disease than *BRCA1* carriers is unclear. Recent evidence suggests that *BRCA1*-related breast cancer is driven by aberrant RANK/RANKL signalling in *BRCA1* heterozygous luminal progenitor cells, coupled with increased DNA damage/defective DNA repair in these cells, resulting in development of basal breast cancers[19]. In contrast, this has not been reported in *BRCA2* carriers, who predominately develop luminal breast cancer. Additionally, *BRCA2*’s predominant reported function is its direct role in homologous recombination-mediated double strand break repair[20]. Clearly, a better understanding of molecular and genetic processes resulting in the development of basal and luminal breast cancers at the single cell level is required. Moreover, given the apparent predominant development of synchronous but distinct cancers in *BRCA2* mutation carriers, the contribution of genomic instability at a single cell level also needs to be investigated. Finally, given recent data demonstrating activation of cell intrinsic innate immune responses to the loss of *BRCA1/2*, the role of early immunoediting in control of tumours in *BRCA1* versus *BRCA2* carriers needs to be investigated[21].

This study is strengthened by relatively complete data. Data for all 211 women from Northern Ireland is complete, with regard to tumour focality, *BRCA* mutation type, age at diagnosis, and tumour type. Furthermore, information about tumour grade, oestrogen receptor status, lymph node involvement, and presence or absence of other primary tumours are all in excess of 96% complete. The considerable quantity of missing HER2 status data reflects the fact HER-2 testing was not routinely carried out at time of diagnosis for many of these women[22]. Additionally, the presence of a large, independent cohort of BRCA1/2 mutation carriers from in the validation cohort provides strong evidence to support the increased prevalence of MF/MC tumours in BRCA2 mutation carriers.

With respect to the long-term outcomes in the Northern Ireland cohort of patients, no difference in outcomes was noted between MF/MC and unifocal tumours, even when adjusted for BRCA mutation status, although this data needs to be interpreted with caution due to the small number of deaths in the two groups. Other groups have reported worse 10 year survival in BRCA1 mutation carriers as compared with BRCA2 carriers, ascribing this difference to tumour biology [23]. However, the cohort of young patients from the POSH study, which form the validation cohort for this study, did not demonstrate a significant difference in overall survival between either BRCA1 or BRCA2 mutation carriers and non-mutation carriers, despite showing a similar increase in multifocality/multicentricity prevalence in BRCA2 mutation carriers [13]. Taken together, these data support the contention that multifocality/multicentricity is not an independent prognostic factor in breast cancer. Due to the retrospective nature of this series, it was not possible to add together the tumour diameters of individual foci as carried out by Fushimi *et al* [2]. Although Fushimi *et al* suggested that doing so may predict for outcome, this may simply be due to the fact that it reflects a higher burden of tumour in these patients rather than being a function of multifocality/multicentricity *per se*.

There are necessarily limitations to a retrospective review of MF/MC breast cancer, as much of the macroscopic pathological information available at the time of initial surgery is no longer available at retrospective slide review. In this study, the assumption was made that the original diagnosis of MF/MC disease (as made by the reporting pathologist with all the macroscopic and microscopic information to hand) was accurate, and a slide review was carried out to determine whether there were any features to suggest that this original diagnosis was incorrect. Indeed in the 10% of cases where slides were reviewed there was no evidence that the diagnosis of MF/MC cancer was incorrect and no cases were excluded on the basis of this review. Furthermore, data on the number of tumour foci and the intervening distance between foci in each case, and their morphological similarities/differences, were not available. Biomarker status (ER/PR/HER2) was also not available for individual tumour foci, as this was generally not assessed on all tumour foci, meaning that it is not possible to comment on the morphological nature of the MF/MC disease in these patients. These limitations are applicable to both the Northern Ireland and POSH study patient cohorts.

In conclusion, we report a higher than anticipated prevalence of multifocality/multicentricity amongst female *BRCA1/2* mutation carriers diagnosed with breast cancer. This finding was seen in a cohort of patients from Northern Ireland, and is validated in the independent cohort of BRCA1/2 mutation carriers from the POSH study. Our data also suggests that multifocality/multicentricity is more common in *BRCA2*-associated breast cancer. Those with MF/MC disease were more likely to be younger at diagnosis, and more likely to be oestrogen receptor positive than those with unifocal disease. These findings have important implications for clinicians involved in the care of patients with *BRCA-*associated breast cancer, who will need to ensure that *BRCA2-*associated tumour focality is thoroughly assessed during the diagnostic process. Furthermore, where breast conserving surgery is being considered as a treatment option for these patients, surgeons need to be aware of the increased incidence of multifocality and plan surgery accordingly, to ensure complete excision at one operation and minimise the consequences associated with re-operation due to involved margins. Finally, further studies are required to establish the underlying mechanistic basis for these findings.

## Data Availability

Data referred to in the manuscript is not publically available as it derives from patient pathology notes and records which are directly identifiable.

## Additional information

### Acknowledgements

Funding for the POSH study has been provided by the Wessex Cancer Trust, Cancer Research UK (C1275/A7572, C22524, A11699, A19187), and Breast Cancer Now (2005Nov53).

SGH is funded by a research fellowship from the Health Education England Genomics Education Programme (HEE GEP).

### Author contributions statement

study idea was conceived by SMcI and KS. Data was collected by ADM, SA, AB and PJM. Data for the POSH cohort was provided by BE, SG-H, ERC, RIC and DME. Statistical analysis was performed by ADM, SA, CM and AB. Verification was performed by PJM and CB. Manuscript was drafted and critically revised by all authors; all authors have approved the final version of the manuscript.

## Table and Figure Legends

**Supplementary Table 1:** Oestrogen receptor status of the Northern Ireland cohort of female BRCA1/2 mutation carriers diagnosed with multifocal breast cancer between 1994-2017. **Pearson’s χ^2^ where p<0.05 indicates significance.

| Women with pathologically confirmed multifocal breast cancer |  | *BRCA1* mutation<br>N (%) | *BRCA2* mutation<br>N (%) | p-value<br>(*) |
| --- | --- | --- | --- | --- |
| Oestrogen receptor status | Positive | 6 (15.4) | 33 (84.6) | 0.039 |
|  | Negative | 6 (50.0) | 6 (50.0) |  |
|  | Missing | 0 (0.0) | 1 (100.0) |  |

**Supplementary Table 2:** Oestrogen receptor status of the POSH study cohort of female BRCA1/2 mutation carriers. Pearson’s χ^2^ where p<0.05 indicates significance.

| Women with pathologically confirmed multifocal breast cancer |  | *BRCA1* mutation<br>N (%) | *BRCA2* mutation<br>N (%) | P value |
| --- | --- | --- | --- | --- |
| Oestrogen receptor status | Positive | 9 (37.5) | 51 (85.0) | <0.001 |
|  | Negative | 15 (71.4) | 6 (28.6) |  |
|  | Missing | 0 (0.0) | 0 (0.0) |  |

## References

1. Vera-Badillo FE, Napoleone M, Ocana A, et al. Effect of multifocality and multicentricity on outcome in early stage breast cancer: a systematic review and meta-analysis. Breast cancer research and treatment 2014; 146: 235–244.

2. Fushimi A, Yoshida A, Yagata H, et al. Prognostic impact of multifocal and multicentric breast cancer versus unifocal breast cancer. Surg Today 2019; 49: 224–230.

3. Rezo A, Dahlstrom J, Shadbolt B, et al. Tumor size and survival in multicentric and multifocal breast cancer. Breast 2011; 20: 259–263.

4. Coombs NJ, Boyages J. Multifocal and multicentric breast cancer: does each focus matter? Journal of clinical oncology : official journal of the American Society of Clinical Oncology 2005; 23: 7497–7502.

5. Neri A, Marrelli D, Megha T, et al. “Clinical significance of multifocal and multicentric breast cancers and choice of surgical treatment: a retrospective study on a series of 1158 cases”. BMC Surg 2015; 15: 1.

6. Kim H, Kim CY, Park KH, et al. Clonality analysis of multifocal ipsilateral breast carcinomas using X-chromosome inactivation patterns. Hum Pathol 2018; 78: 106–114.

7. Noguchi S, Aihara T, Koyama H, et al. Discrimination between multicentric and multifocal carcinomas of the breast through clonal analysis. Cancer 1994; 74: 872–877.

8. Eeles R, Knee G, Jhavar S, et al. Multicentric breast cancer: clonality and prognostic studies. Breast cancer research and treatment 2011; 129: 703–716.

9. Desmedt C, Fumagalli D, Pietri E, et al. Uncovering the genomic heterogeneity of multifocal breast cancer. The Journal of pathology 2015; 236: 457–466.

10. Kanumuri P, Hayse B, Killelea BK, et al. Characteristics of Multifocal and Multicentric Breast Cancers. Annals of surgical oncology 2015; 22: 2475–2482.

11. Venkitaraman AR. Functions of BRCA1 and BRCA2 in the biological response to DNA damage. J Cell Sci 2001; 114: 3591–3598.

12. Eccles D, Gerty S, Simmonds P, et al. Prospective study of Outcomes in Sporadic versus Hereditary breast cancer (POSH): study protocol. BMC Cancer 2007; 7: 160.

13. Copson ER, Maishman TC, Tapper WJ, et al. Germline BRCA mutation and outcome in young-onset breast cancer (POSH): a prospective cohort study. The lancet oncology 2018; 19: 169–180.

14. Bursac Z, Gauss CH, Williams DK, et al. Purposeful selection of variables in logistic regression. Source Code Biol Med 2008; 3: 17.

15. Mavaddat N, Barrowdale D, Andrulis IL, et al. Pathology of breast and ovarian cancers among BRCA1 and BRCA2 mutation carriers: results from the Consortium of Investigators of Modifiers of BRCA1/2 (CIMBA). Cancer epidemiology, biomarkers & prevention : a publication of the American Association for Cancer Research, cosponsored by the American Society of Preventive Oncology 2012; 21: 134–147.

16. Bergthorsson JT, Ejlertsen B, Olsen JH, et al. BRCA1 and BRCA2 mutation status and cancer family history of Danish women affected with multifocal or bilateral breast cancer at a young age. Journal of medical genetics 2001; 38: 361–368.

17. Foulkes WD, Metcalfe K, Sun P, et al. Estrogen receptor status in BRCA1- and BRCA2-related breast cancer: the influence of age, grade, and histological type. Clinical cancer research : an official journal of the American Association for Cancer Research 2004; 10: 2029–2034.

18. Wang L, Di LJ. BRCA1 and estrogen/estrogen receptor in breast cancer: where they interact? Int J Biol Sci 2014; 10: 566–575.

19. Nolan E, Vaillant F, Branstetter D, et al. RANK ligand as a potential target for breast cancer prevention in BRCA1-mutation carriers. Nat Med 2016; 22: 933–939.

20. Orr KS, Savage KI. The BRCA1 and BRCA2 Breast and Ovarian Cancer Susceptibility Genes — Implications for DNA Damage Response, DNA Repair and Cancer Therapy. In. (ed)^(eds). InTech, 2015.

21. Parkes EE, Walker SM, Taggart LE, et al. Activation of STING-Dependent Innate Immune Signaling By S-Phase-Specific DNA Damage in Breast Cancer. Journal of the National Cancer Institute 2017; 109.

22. Dowsett M, Hanby AM, Laing R, et al. HER2 testing in the UK: consensus from a national consultation. J Clin Pathol 2007; 60: 685–689.

23. Soenderstrup IMH, Laenkholm AV, Jensen MB, et al. Clinical and molecular characterization of BRCA-associated breast cancer: results from the DBCG. Acta oncologica (Stockholm, Sweden) 2018; 57: 95–101.

